# Auditing What Was Said: The Epistemic Promise and Limits of Ambient AI in Clinical Practice

**DOI:** 10.64898/2026.04.29.26351997

**Authors:** Vincent Misrai, Alena Bruchon, Alexis Campan, Jean-Michel Loubes, Antoine Piau

## Abstract

Traditional audit methods that rely on written records often miss the nuances of clinical reasoning that influence patient care. Ambient artificial intelligence captures spoken clinical encounters, allowing the analysis of real clinician–patient dialogue at scale. In a study of 124 urology consultations, a transcript-centered audit identified inter-physician variation and expert disagreement that conventional review missed. We explore the epistemic gains of this approach, its nonverbal blind spots, behavioral effects, technical vulnerabilities, and the EU AI Act’s regulatory landscape.

## The Epistemic Shift

Consider a 68-year-old patient newly diagnosed with prostate cancer who receives conflicting recommendations from two urologists. Both consultations are thoroughly documented and guideline concordant. Yet ambient AI transcripts reveal a critical distinction: one clinician explicitly elicits and incorporates the patient’s values regarding sexual function and continence before recommending a course of action, whereas the other does not. A conventional audit limited to written records would treat both encounters as equivalent. A transcript-centered audit makes this difference visible, enabling rigorous evaluation of the transparency and completeness of the clinical reasoning shared with the patient, not just the final diagnostic label (Figure 1).

**Figure 1.**
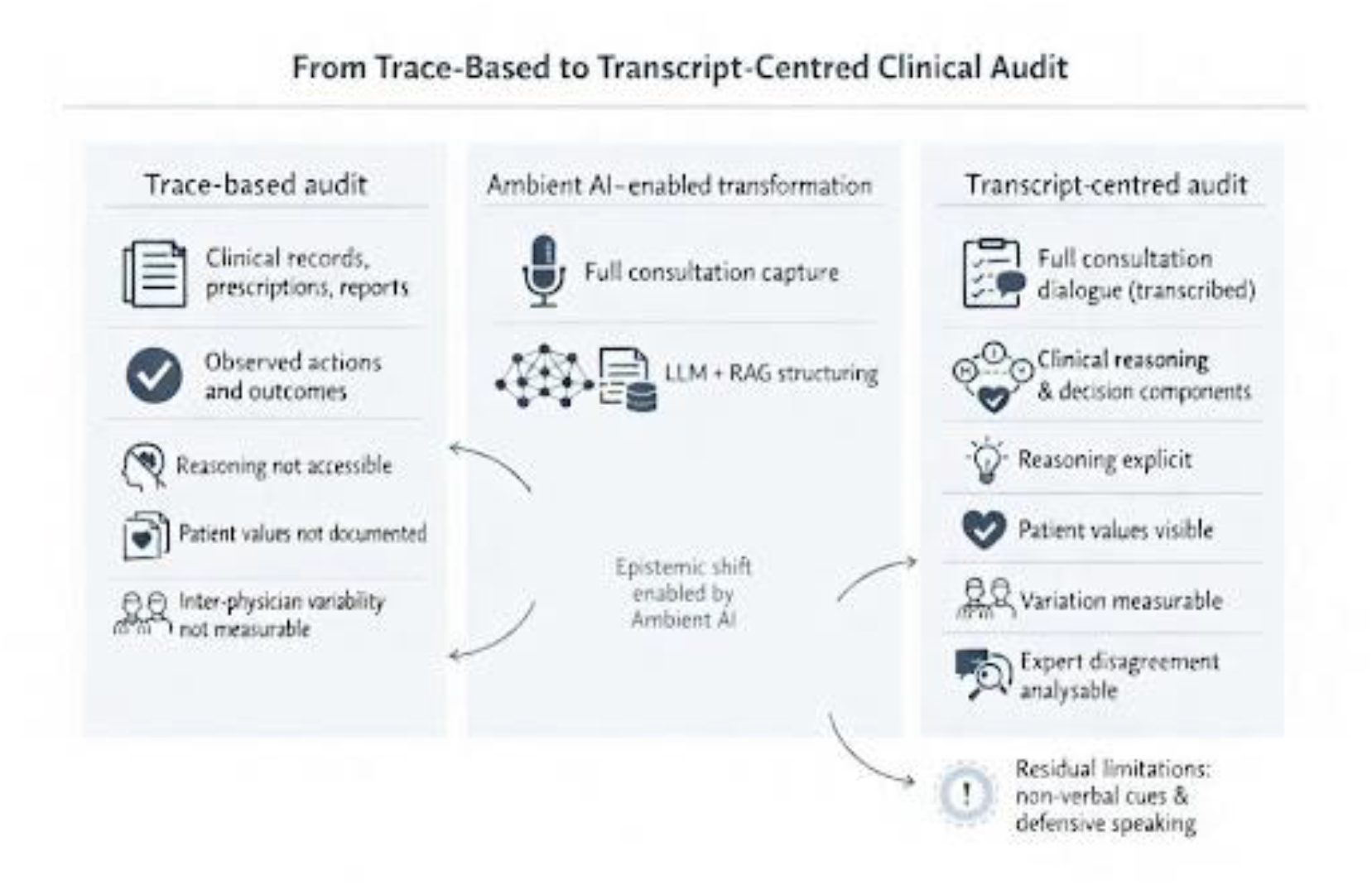
From Trace-Based to Transcript-Centered Clinical Audit. The left panel shows the epistemic limits of traditional trace-based audit: written records capture actions and outcomes but do not reveal clinical reasoning, patient values, or inter-physician differences. The centre panel highlights three AI capabilities: scalable consultation capture, structured extraction by large language models, and retrieval-augmented generation based on clinical guidelines. These enable the epistemic shift, acknowledging the residual blind spots of audio capture and vulnerabilities of speech recognition. The right panel illustrates what transcript-centered audit reveals: verbalized clinical reasoning, expressed patient values and trade-offs, measurable physician variation, and divergences in evaluative thresholds among reviewers, which traditional audit suppresses.

### Persistent Limitations of Trace-Based Audit

Audit and feedback are established tools for reducing unwarranted clinical variation. ^1,2^However, their effects have remained modest across specialties and settings. ^3^ Physician resistance and logistical burden are commonly cited explanations, but a more fundamental issue persists.

Clinical deviation (underuse, overuse, or misuse of care) drives practice variation. ^4^ Conventional audit detects deviation by examining records, prescriptions, and reports. These artifacts document outcomes but do not capture the decision-making process. They rarely reveal how clinicians weigh evidence, handle uncertainty, or negotiate with patients.

This gap has structural consequences. Audit systems that assess only documented outputs cannot access the reasoning behind clinical decisions. Discrepancies may be flagged, but their causes remain opaque. When clinicians act as both authors and interpreters of their own records, the process becomes circular, often favouring selective rather than reflective documentation. The operative target of improvement becomes the record, not the reasoning.

Trace-based audit will therefore always operate under a structural ceiling. Addressing this limitation requires a different primary data source, which ambient AI now provides. Routine recording of consultations yields information that extends beyond documentation: it allows direct observation of diagnostic hypotheses, the options discussed, the concerns raised, and the values articulated by patients. The shift from written record to recorded dialogue is not merely a technical upgrade.^5^ It is a qualitative change in what can be known and measured about clinical care. Audit can then address reasoning as it unfolded during the encounter, rather than inferring it from its documentary residue.

### The Non-Verbal Epistemic Gap

The limits of this approach must be acknowledged. Audio transcripts capture only what is spoken and omit visual cues, nonverbal communication, and physical examination findings that are not verbalized. If a clinician’s decision is influenced by a patient’s visible frailty that is not articulated aloud, a transcript-based audit may wrongly judge the reasoning as incomplete.

Transcripts offer a partial window into clinical reasoning. Recent work has also shown that probabilistic ambient scribes can omit or fabricate details, further constraining their reliability.^8^ A transcript-centered audit expands what is observable but does not provide a complete or infallible account.

### Defensive Speaking: A New Hawthorne Effect

Trace-based auditing has long encouraged selective documentation. A transcript-centered audit, by focusing on spoken interactions, may prompt a different behavioral shift. Clinicians who know their words will be evaluated against guidelines may begin to speak defensively: verbalizing negative findings, reciting rationales, or justifying decisions that would otherwise remain unspoken. This defensive speaking can disrupt the natural flow and empathic quality of the encounter and may diminish communication over time.^7^ Audit systems should therefore minimize intrusiveness and clearly separate governance from individual performance sanctions.

### Architectural Requirements

Realizing ambient AI’s analytical potential requires three capabilities working together. First, ambient AI systems enable scalable, auditable data capture embedded in routine clinical practice.^6^ Second, large language models extract decision-relevant structure from unstructured dialogue.

Third, retrieval-augmented generation (RAG) grounds evaluations in explicitly retrieved guideline passages, making the basis of every audit flag transparent and contestable.^9^ Together, these capabilities enable normative comparison at a scale human reviewers alone cannot sustain

To operationalise these capabilities, we developed RAGtime, an in-house hybrid retrieval-augmented generation system. RAGtime orchestrates a large language model over a multi-stage retrieval pipeline grounded in the 2025 editions of the French (AFU) and European (EAU) urological guidelines. The system flags candidate deviations from these guidelines and classifies them as warranted or unwarranted, depending on whether the transcript contains an explicit clinical justification. Each unwarranted deviation is then graded along two independent axes — the normative strength of the violated recommendation and the anticipated patient-level impact. The full technical configuration of the pipeline, the expert review protocol, and the statistical analyses are detailed in the Methods section below. Internal components — chunking, graph schema, retrieval and re-ranking parameters, prompt templates, and agentic orchestration — remain proprietary.

A further architectural requirement specific to retrospective audit warrants emphasis: temporal version control of the guideline corpus.^1^} Clinical guidelines are updated regularly, but audited consultations occurred at a specific point in time. A vector store populated with a later version of a guideline could, if used to audit earlier consultations, incorrectly flag decisions that were appropriate at the time. Audit systems must therefore track guideline versions, specify the validity period of each, and retrieve only passages in force on the date of the consultation. This requirement did not arise in the present pilot, as both the audited consultations and the guideline editions were from 2025. It becomes essential whenever audits span several years.

## Methods

### Study design

This work reports the baseline phase (run 1) of a prospective, single-centre audit-and-feedback cycle evaluating the integration of transcript-centred clinical audit into routine urological practice. The complete cycle comprises two consecutive runs, separated by an academic review and by structured feedback to participating physicians. Run 1 covered consultations audited between November 2025 and January 2026; academic review and adjudication followed in February 2026; individualised feedback was restituted to participating physicians in early March 2026; and run 2 is currently nearing completion. The present manuscript reports run 1 and the methodology of the audit framework. The comparative analysis of run 2 against run 1 — designed to evaluate whether audit-and-feedback modifies physician behaviour — will be the subject of a separate report.

### Setting and participants

Seven practising urologists at a tertiary urological centre (Clinique Pasteur, Toulouse, France) recorded all consecutive outpatient consultations between November 2025 and January 2026 using the deployed ambient AI scribe. All such consultations were eligible for inclusion. The final cohort comprised 124 consultations, distributed unevenly across physicians (median 17 consultations per physician; range 11 to 32), reflecting differences in clinical activity during the inclusion period. No consultations were excluded on the basis of complaint type, duration, or patient demographics. The cohort spanned the full breadth of urological practice, including urological oncology, lower urinary tract symptoms and bladder dysfunction, urolithiasis, andrology, and follow-up encounters.

### Data acquisition and privacy-preserving pipeline

Consultations were documented using Nabla Copilot, an ambient AI scribe deployed in routine clinical practice at the study site for the primary purpose of clinical documentation. The scribe transcribed the clinician–patient dialogue in real time and produced an initial structured clinical note. Audio streams were destroyed by the vendor immediately after transcription; no audio was retained or accessible to the research team at any point. Transcripts were retained by the vendor on a 15-day rolling window. The attending physician reviewed and corrected the AI-generated draft to produce a finalised clinical report, which, alongside the corresponding raw transcript, served as the primary data source for the audit. Before ingestion by the audit pipeline, two trained clinical research associates manually pseudonymised the transcripts and corrected clinical reports. They systematically suppressed direct patient identifiers and incidental third-party information — references to family members, accompanying persons, or third-party medical history mentioned during the encounter.

### Audit pipeline

The audit relied on an in-house hybrid retrieval-augmented generation system (RAGtime). RAGtime orchestrated a large language model (Gemini 3 Pro) over a multi-stage retrieval pipeline that combined dense semantic search, a neural re-ranking layer, and graph-based contextual analysis. The reference corpus comprised the 2025 editions of the French (AFU) and European (EAU) urological guidelines. Guideline passages were embedded using PubMedBERT, a transformer pre-trained on biomedical text, and indexed in a vector store. A knowledge graph implemented in Neo4j encoded the structured clinical reasoning of the guidelines — diagnostic pathways, treatment recommendations with their conditions and contraindications, and cross-specialty considerations. The graph then provided a downstream contextualisation layer over the re-ranked candidate passages. For each consultation, the pipeline performed entity extraction from the pseudonymised transcript and clinical report, hybrid retrieval over the indexed corpus, and grounded adjudication. The output was a structured list of candidate deviations. Each deviation received two independent grades: a normative-strength grade derived from the source recommendation, and a clinical-impact grade assessed separately. The internal chunking strategy, graph schema, retrieval and re-ranking parameters, prompt templates, agentic orchestration, generation parameters, and grade-mapping rules remain proprietary and are not disclosed.

### Expert review protocol

Three senior academic urologists served as expert reviewers. The panel was assembled to cover the full urological scope encountered in the cohort, spanning oncological, functional, and onco-functional domains. All three reviewers actively contribute to the development of national and European urological guidelines. The panel auditing the system is therefore composed of authors of the normative corpus that the system applies. For each of the 124 consultations, each reviewer independently audited the system output against the pseudonymised transcript and the corrected clinical report. Reviewers were blinded to physician identity but not to system outputs. This was a deliberate methodological choice. Raw automatic speech recognition transcripts contain noise — disfluencies, mistranscriptions, overlapping speech — that makes fully blinded transcript-only review impractical. Conversely, review based solely on the corrected clinical report would have defeated the central premise of the framework by reverting to documentary trace. The chosen protocol therefore asked reviewers to verify each system-flagged deviation against the transcript and clinical report, and to independently scan the same materials for deviations the system may have missed. We acknowledge that this design favours confirmation of system outputs over detection of false negatives, and we treat reported sensitivity as an upper-bound estimate.

Review was structured by chief complaint. Each reviewer first identified the motif of the consultation and then assessed the encounter against the elements they considered essential for that motif. In suspected prostate cancer, for instance, this meant checking whether all therapeutic options had been presented and whether the patient’s values regarding functional outcomes had been elicited. This pattern-based approach reflects how senior academic urologists deploy expertise in real-world peer review. We deliberately chose not to enforce a single standardised checklist across reviewers; the resulting variability in evaluative thresholds is itself an object of study and is reported in the inter-rater agreement analysis. A deviation was officially confirmed when at least two of the three reviewers endorsed it. For each consensus-confirmed deviation, reviewers then assigned a binary impact rating, low or high. High impact was assigned when the reviewer judged that the deviation could reasonably lead to severe complications or contribute to mortality, taking into account the patient’s comorbidity profile, age, frailty, and the causal pathways plausibly initiated by the deviation. This judgement was made independently per deviation, since a single consultation could involve several deviations with distinct impact profiles. High impact was retained only on unanimous agreement among the three reviewers.

### Statistical analysis

All analyses were performed in Python (pandas, NumPy, SciPy, scikit-learn). We quantified inter-rater agreement at three complementary levels. At the pairwise level, we computed raw agreement, Cohen’s κ, and the Jaccard index for each reviewer pair, with 95% confidence intervals derived from non-parametric row-level bootstrap (20 000 resamples). At the three-rater level, we used Fleiss’ κ with bootstrap 95% CI, and we reported the proportion of unanimous ratings with Clopper–Pearson exact 95% CI. We tested systematic bias between reviewer pairs with exact McNemar tests on discordant pairs, and global homogeneity across the three reviewers with Cochran’s Q. System performance against the consensus reference standard was summarised by accuracy, precision, recall (sensitivity), specificity, negative predictive value, F1, and false-positive and false-negative rates. Proportion-based metrics carry Clopper–Pearson exact 95% CI. As argued above, these metrics indicate system–panel concordance under conditions of slight-to-fair inter-rater agreement; they are not estimates of an objective clinical truth. We used a two-sided α of 0.05 throughout. The random seed for all bootstrap procedures was fixed (12 345) to ensure reproducibility. Analysis code is available from the corresponding author upon reasonable request.

### Illustrative Proof-of-Concept

To close this examination, we evaluated the integration of transcript-centred audit into routine clinical workflow. The aim was to determine whether the theoretical and technical advances described above could deliver meaningful, actionable information when applied to real-world data from 124 consecutive urology consultations at a single academic centre.

Three independent academic urologists reviewed each AI-generated audit output against both the raw transcript and the validated clinical report. A majority-consensus rule was applied (≥2 of 3 reviewers), with unanimous agreement required for findings of high patient impact. Across 124 consultations, the system produced 156 audit outputs; expert consensus confirmed 81 deviations, of which 70 were classified as unwarranted. Outputs indicating no deviation were confirmed in 96% of cases, while positive flags were endorsed in 74%. Only 36% of confirmed deviations achieved unanimous agreement, with the remainder requiring majority arbitration. Most discordant cases were false positives, indicating a systematic tendency toward over-signalling.

Two findings carry implications beyond the pilot context. First, expert-confirmed deviation rates ranged from 0.5 to 0.9 per consultation across participating physicians, a nearly twofold variation that conventional documentation-based audit of the same encounters would not have detected.

Second, expert reviewers disagreed substantially on which outputs constituted meaningful deviations. Inter-rater agreement, measured by Fleiss’ κ, was only slight to fair, a level consistent with prior studies of peer review among specialist physicians.^11,12^This level of disagreement persisted despite all three reviewers being academic urologists applying identical guideline references to the same transcript evidence. Such divergence suggests that variation in evaluative thresholds is intrinsic to clinical judgement rather than attributable to differences in training or information access.

For each confirmed unwarranted deviation, we evaluated both the normative strength of the violated recommendation (based on guideline grading) and the anticipated patient-level impact (by majority consensus). Impact was assessed per deviation, recognising that a single consultation can involve multiple deviations with distinct profiles. The two axes proved empirically dissociated: among 28 confirmed unwarranted deviations from strong recommendations, only 5 carried high anticipated impact, whereas 20 carried low impact.

Omitting a validated symptom score in benign prostatic hyperplasia, for instance, violates a strong recommendation but carries no prognostic weight. Conversely, some violations of lower-strength recommendations were associated with substantial anticipated harm. Prescribing an anticholinergic to a nonagenarian, for example, may initiate a causal chain of delirium, falls, and hip fracture, with one-year mortality approaching 30%. Audit systems that collapse normative strength and clinical consequence into a single grade risk misdirect attention; a two-dimensional grading framework is essential.

### The Kappa Paradox and the Risk of Alert Fatigue

Two methodological caveats are important for interpreting these results. First, the ground truth for sensitivity and precision is inherently unstable when reviewer agreement is only slight to fair. Reporting precise performance metrics in this context illustrates the so-called kappa paradox: the system’s apparent accuracy reflects how closely it matches the average bias of the reviewer panel rather than an objective clinical truth. This mirrors long-standing findings from medical-record review, where adverse-event detection is reproducible, but judgements of preventability are not.^11,12^Headline metrics from this pilot should therefore be viewed as indicators of system–panel concordance, not as definitive measures of performance.

Second, a system that persistently over-signals will generate alert fatigue at scale and risks disengagement from the very clinicians it aims to support. Effective deployment will require careful tuning of evaluative thresholds to improve specificity, calibration with more diverse reviewer panels, and operating modes that prioritise high-impact deviations over exhaustive flagging. Low inter-rater agreement does not invalidate the framework; it exposes real-world variability that conventional audit, often conducted by a single reviewer on self-authored records, tends to obscure. Transcript-centred audit brings this variability to light, making it measurable and actionable.

These observations require further investigation. The single-centre, single-specialty design and the absence of a fully blinded review protocol limit generalisability. The inter-physician variation reported here should therefore be interpreted as an observation requiring confirmation under fully blinded multi-center conditions, rather than as a population-level estimate.

### Conditions of Legitimate Implementation

Technical feasibility does not ensure legitimate adoption. The legal architecture supporting this approach must be addressed explicitly. An audio recording of a medical consultation captures voice, and voice qualifies as biometric data under Article 9 of the General Data Protection Regulation when it is persistently stored. This triggers heightened protection. The framework adopted in this pilot circumvents the exposure by leveraging a privacy-by-design architecture already embedded in the deployed ambient AI scribe. Audio streams are transcribed in real time and immediately destroyed by the vendor, so that no audio is ever retained or processed by the research team. The legal basis for the secondary research use of the resulting transcripts and clinical reports — pseudonymised manually before any analysis — is the French CNIL MR-004 reference methodology, which applies to retrospective research on documentary outputs of routine clinical care. This design preserves full patient autonomy through documented non-opposition, which may be withdrawn at any time without consequence for care. It also avoids the more onerous explicit opt-in consent that would otherwise apply to the prospective placement of dedicated recording devices for research. Future deployments that retain audio — for instance, to study prosody, paralinguistic markers, or speech-recognition equity — would fall outside this framework and would require explicit Article 9 consent.

Transcript-centred audit must also be clearly distinguished from a second medical opinion. A second opinion is a prospective therapeutic act: it creates a therapeutic relationship and entails professional liability. Audit, by contrast, is retrospective and pedagogical. It evaluates whether the reasoning documented in the transcript was adequately supported and guideline-concordant at the moment of decision, not whether the outcome was ultimately correct. This temporal boundary is central to the framework’s medicolegal positioning.

Retrospective use mitigates, but does not eliminate, regulatory exposure. While such systems fall outside the scope of real-time Software as a Medical Device classifications under EU MDR, they remain governed by the EU AI Act.^13^ Because they evaluate the work performance and professional conduct of identifiable individuals, AI systems used to audit physician practice are categorically classified as high-risk under Annex III of that regulation. Anticipating the corresponding obligations regarding transparency, human oversight, robustness, and data governance is integral to legitimate implementation.

A further data-protection imperative concerns third parties. Free-flowing clinical dialogue routinely captures sensitive information about persons other than the index patient: a relative’s oncological history mentioned in passing, a spouse’s psychiatric treatment, or the contributions of an accompanying family member. None of these third parties has consented to participate in an audited recording. Data-minimisation pipelines must therefore be engineered to detect and sanitise bystander information before transcripts are stored persistently or subjected to secondary analysis. Relevant techniques include automated named-entity recognition, role-based diarisation, and the suppression or pseudonymisation of third-party utterances. Without such safeguards, the framework extends the consultation’s privacy footprint well beyond the boundaries of the consenting patient.

A complementary equity concern arises upstream of the auditor itself. Transcript-centred audit depends on automated speech recognition, which exhibits well-documented degradation in word error rate for non-native accents, regional dialects, low-frequency vocabularies, and the dysarthric or presbyphonic voices common in elderly patients. A clinician who explicitly verbalises a contextual justification but is mistranscribed will be penalised by the downstream RAG auditor for an apparently unwarranted deviation. Conversely, mistranscribed patient utterances may erase the very values and preferences the framework purports to render visible. Left unaddressed, the pipeline risks institutionalising sociolinguistic bias under the appearance of algorithmic neutrality. Continuous algorithmic-equity audits must therefore be a standing requirement of any deployment, not an optional enhancement. Such audits should stratify ASR performance and downstream audit outcomes by clinician and patient sociolinguistic characteristics, and trigger recalibration whenever disparities exceed predefined thresholds.^8^

Patient consent must be explicit, obtained in advance, and remain revocable without consequence for care. Clinicians should have enforceable control over how audit outputs are used and accessed. A system perceived as a surveillance tool will provoke the same defensive behaviours observed with trace-based audit and further entrench defensive speaking.

Institutions should anticipate and accommodate clinician disagreement with audit findings. Guidelines are grounded in population-level evidence, whereas individual clinical decisions are shaped by contexts that no guideline can fully capture. Trace-based audit is particularly vulnerable to these objections because the auditor lacks access to the underlying reasoning and context. Transcript-centred audit addresses many of these limitations by preserving the full clinical dialogue, enabling both auditor and clinician to operate on the same evidential basis.

When disagreements persist, they shift toward how population-level recommendations apply to individual cases, a more constructive and clinically relevant debate than disputes over incomplete records.

Mechanisms that allow clinicians to contest or contextualize audit findings are essential. These safeguards do not merely protect against evidentiary asymmetry; they are the process by which the applicability of population-level recommendations to individual cases is progressively refined.

### An Epistemic Inflection Point

Ambient AI does not eliminate the core interpretive challenge of clinical audit. Our proof-of-concept evaluation showed that even experienced specialists, working from the same guidelines and reviewing identical consultations, disagreed on what constituted a deviation. This interpretive variability is not a flaw to be engineered away; it is a central reality that audit systems must be designed to surface and address.

Transcript-centred audit fundamentally shifts the focus of clinical evaluation. Rather than verifying documentary compliance, it renders the reasoning behind clinical decisions explicit, structured, and open to scrutiny. It also recognises the non-verbal elements that remain beyond reach and the behavioural adaptations that may follow its adoption. By making inter-physician variation measurable and clarifying evaluative thresholds, this approach transforms disagreement from a problem to be concealed into a productive part of quality improvement.

The key question is no longer whether clinical dialogue can be recorded, but whether healthcare systems will use this capability to assess clinical reasoning or simply to automate record-keeping. If ambient AI is used only for documentation, its impact on audit practice will be incremental at best. If it is harnessed to evaluate the reasoning, justifications, and trade-offs underlying clinical decisions within a transparent, consented, and auditable governance framework, it can render what has long remained structurally unobservable observable. Audit and feedback may then begin to address what has eluded trace-based approaches for decades.

## Author Contributions

V.M. conceived the study and drafted the manuscript. A.B. and A.C. contributed to the technical design and implementation of the framework. J.-M.L. contributed to methodological development, analysis, and interpretation. A.P. contributed to study design, clinical interpretation, and critical revision of the manuscript. All authors reviewed and approved the final version.

## Funding Declaration

No funding

### Competing Interests

Drs. Misrai and Piau, Ms. Bruchon, and M. Campan report equity ownership in a company developing AI-based tools for clinical audit and quality improvement. Dr. Loubes reports no financial interests but is involved in methodological work on transparency and explainability for AI-based audit and feedback systems.

### Ethics Statement

The pilot evaluation was reviewed and granted a favorable ethics opinion (avis favorable) by the Comité d’Éthique de la Recherche en Urologie (CERU) of the Association Française d’Urologie (AFU), Maison de l’Urologie, 11 rue Viète, 75017 Paris, France, on 26 May 2025 (submission reference: CERU_2025013; protocol title: “RAGTIME — Retrieval-Augmented Generation To Improve Medical Evaluation”; principal investigator: Vincent Misrai; study site: Clinique Pasteur, Toulouse, France).

Clinical consultations were documented using Nabla Copilot, an ambient AI scribe deployed in routine clinical practice at the study site. Audio streams were transcribed in real time and immediately destroyed by the scribe vendor; no audio recording was retained or accessible to the research team. Resulting transcripts were retained by the vendor on a 15-day rolling window before deletion. Patients were informed of the use of the ambient AI scribe for clinical documentation and of the potential secondary research use of resulting transcripts and validated clinical reports for quality-of-care audit purposes; all patients signed a non-opposition form covering both the clinical use of the scribe and the secondary research use of resulting documents, and all signed forms have been retained by the investigator. Patients could withdraw their non-opposition at any time without consequence for care.

Prior to any analysis, transcripts and corrected clinical reports were pseudonymised manually by two trained clinical research associates, with systematic suppression of direct identifiers and of incidental third-party information. The retrospective secondary use of these pseudonymised documents complied with the French CNIL MR-004 reference methodology (Délibération n° 2018-155, 3 May 2018). As no audio data were retained or processed by the research team, the present study did not entail processing of biometric data within the meaning of Article 9 of the General Data Protection Regulation.

### Data Availability

The anonymized data supporting the findings of this study are available from the corresponding author upon reasonable request, subject to applicable data-protection constraints.

### Code Availability

The full source code of the agentic RAG pipeline is proprietary and not publicly available. In line with Nature Portfolio’s open-science expectations, the methodology, the public components of the technical stack, and the boundary between disclosed and proprietary elements are described in the Methods section. Statistical analysis scripts are available from the corresponding author upon reasonable request.

## Notes

### Funding Statement

This study did not receive any funding

### Author Declarations

The pilot evaluation was reviewed and granted a favorable ethics opinion (avis favorable) by the Comité d’Éthique de la Recherche en Urologie (CERU) of the Association Française d'Urologie (AFU), Maison de l’Urologie, 11 rue Viéte, 75017 Paris, France, on 26 May 2025 (submission reference: CERU_2025013). Participating patients provided explicit written opt-in consent for the audio recording of their consultation, in compliance with the General Data Protection Regulation and applicable French CNIL guidance on the processing of biometric and health data. The retrospective secondary use of resulting transcripts and validated clinical reports relied additionally on the non-opposition framework according to the CNIL methodology. Consent could be withdrawn at any time without consequence for care.

